# Changes in Manual Therapy Techniques from the Perspective of Physical Therapists

**DOI:** 10.1101/2022.09.02.22279416

**Authors:** Nelson A. Rojas, Krista M. Jones, Tami Goetz, Shikha Prashad

## Abstract

**Objective:** Prior research has focused on comparing the effects of manual therapy techniques used in physical therapy in different conditions to determine which technique is more effective and less invasive for the patient. However, no focus has been placed on identifying and evaluating the changes in manual therapy techniques. This study aims to evaluate the changes in manual therapy techniques from the perspective of current and retired physical therapists who are or were licensed in Washington state.

**Methods:** Eighteen participants completed a modified Physical Therapy Profile Questionnaire (PTPQ) to explore their experience with manual therapy techniques and whether they implemented changes in their practice of these techniques.

**Results:** The results suggest that changes in manual therapy techniques occur as physical therapists gain and develop more experience working with many clients and treating different conditions. Joint mobilization, taping, and muscle energy techniques were the most used and recommended manual therapy techniques by physical therapists in this sample. There was no significant correlation between the years of practice and the number of manual therapy techniques selected by participants (r = -0.044, p = 0.887).

**Conclusions:** This study suggests that changes and adjustments in manual therapy techniques have occurred throughout time as physical therapists gain practice in the field and can identify what manual technique will produce a better patient outcome in a specific patient.

**Impact Statement:** This study examined how the use of manual therapy techniques has evolved in field of the physical therapy and the factors that contribute to these changes.

## Introduction

Physical therapy is a long-changing profession that has expanded the possibilities of treatment for individuals recovering from an acute injury, surgery, or medical condition. The field continues to change to advance clinical practices and technologies and improve clinical outcomes. These advances allow for the modification of therapy interventions and the evaluation of techniques. There have been many changes in the implementation of physical therapy since the foundation of the American Physical Therapy Association in 1921. One of the biggest changes has been in manual therapy, which is a procedure that specifically targets musculoskeletal regions of the body to improve musculoskeletal function and reduce pain levels^1,2^. Manual therapy techniques such as myofascial release (a combination of sustained pressure movements to stretch and lengthen the muscle fascia), muscle energy techniques (mobilization of joints and stretching of tight muscles and fascia), thrust versus non-thrust manipulation (skilled passive movements to joints and/or related soft tissues at varying speeds and amplitudes), dry needling (treating underlying muscular trigger points to restore neuromusculoskeletal function), Mulligan’s technique (combination of natural and sustained apophyseal glides and mobilization with movement to treat musculoskeletal injuries), and soft tissue mobilization techniques (combination of manual techniques that use stretching and deep pressure to improve musculoskeletal function) have revolutionized the rehabilitation field^3^. Modifications and adjustments occur over time as new research guides therapists towards safer and more efficient options to treat patients. Although many changes in manual techniques have taken place, to our knowledge, no studies have systematically identified the changes in this universally used method in physical therapy.

Manual therapy practice references can be traced back to 400 BCE^4^. The practice of spinal manipulation was highly utilized in many ancient western civilizations and remote world communities^4^. Hippocrates initially described spinal manipulation in his books as the use of gravitational forces to treat musculoskeletal conditions^4–6^. Additionally, Hippocrates described the setting of joints by the method of leverage, a similar method of joint mobilization used in practice currently. He recommended the use of this manipulation therapy as well as the manipulation of the spine through prone traction^5^. Through his work, Hippocrates demonstrated the effectiveness of these therapy techniques and recommended modifications by manipulating the different parameters (i.e., speed, direction, and frequency of the manipulations) with the inclusion of high-velocity thrust manipulation techniques^6^. Furthermore, C. Galen, a surgeon who lived from 131 to 202 CE, applied manual therapy to the spine and extremities of the body through joint mobilization and massage^5^. Galen also utilized the manual techniques introduced by Hippocrates, some of which are still included in the medical literature. For over 1600 years, the manipulation methods and treatment designs of Hippocrates were widely used and referenced in research and medical texts^4^. Massage therapy is the earliest form and most practiced manual therapy technique. Reports from 1584 indicate the delivery of lectures at Cambridge University about massage, exercise, and hydrotherapy^4^. Since then, massage therapy has continued to expand and gain popularity. By the early 20th century, physical therapists were trained and performed spine manipulation as these therapeutic practices became more widely accepted^6^. Physical therapy was formally founded in England in 1899 and was officially established in the United States in 1921^5,7^.

Early studies aimed to critically assess the role of manual therapy within the profession of physical therapy. As an emerging field, Farrell, and Jensen^8^, posed several questions, including which treatments would collaborate best with patients and what allows patients to get better. This initial work influenced others like Fitzgerald et al. to identify potential problems in evaluating the effectiveness of this new manual therapy field^9^. Fitzgerald et al. suggested that clinicians perform more experimental studies due to the lack of knowledge of the effects of different treatment techniques, reliable and valid measures, progression of treatments, and treatment effectiveness. More recently, new research has attempted to answer these questions by comparing multiple manual therapy techniques, their mechanisms, and their effectiveness. In a study conducted by Hoeksma et al., manual therapy had a higher success rate in treating patients with osteoarthritis of the hip compared to an exercise therapy program^10^. This study suggested that the group of patients who underwent manual therapy had significantly better outcomes in overall hip function, range of motion, pain, and stiffness compared to the exercise therapy group. Furthermore, a study conducted by Kachingwe et al. compared the effectiveness of manual therapy techniques and an exercise program in the treatment of shoulder impingement^11^. The findings from this study were similar to those found by Hoeksma et al.^10^, but suggested that for this specific condition, the combination of mobilizations and an exercise program resulted in better patient outcomes. The results from these studies indicated that manual therapy is an effective method for therapists to treat patients. However, these studies also suggested that future studies should compare other manual therapy techniques to evaluate their relative effectiveness.

In more recent years, there has been an increase in studies that evaluate and compare newer and older manual techniques to identify which are more effective in practice. One of these evaluated techniques is constraint-induced movement therapy that aims to treat individuals who have suffered from a cerebrovascular accident. Constraint-induced movement therapy helps improve upper-limb function in individuals who have suffered from stroke or any other central nervous system damage by constraining them to use their affected limb while the non-affected side is restrained. Numerous studies have shown that this technique is an effective treatment that produces big improvements^12–16^ but it was unclear why this technique is effective^17,18^. Taub et al. suggested that this form of therapy is effective due to the intensive training of the affected limb while constraining the non-affected limb^19^. In a later study, Taub et al.^20^ evaluated constraint-induced movement therapy in combination with some conventional neurorehabilitation techniques in patients with plegic hands (i.e., paralysis of the hand in which all voluntary muscular movement is lost) caused by a stroke. They found that patients with a plegic hand benefited from constraint-induced movement therapy in combination with some conventional rehabilitation techniques despite having a brain injury^20^. The authors hypothesized that adequate methods could stimulate and help rewire neuroplastic changes and unveil restrained motor capacity to generate therapeutic effects in patients who have suffered from severe brain injury. Although constraint-induced movement therapy has been effective, new modified versions of this therapy have been even more effective without the need of constraining patients for lengthy periods of time, a key disadvantage of the traditional form. Bang et al. investigated the modified version of constraint-induced movement therapy that consisted of only restraining the least affected side for about five hours of the day while the more affected side received intense and repetitive training for up to two hours^21^. They also combined the modified version with a strap that was attached around the thoracic vertebral level of the participants to the back of a chair to limit any compensatory trunk movements (e.g., rotation of the trunk or sagittal displacement). The authors found that the modified version was more effective in improving the overall function in the upper extremities in subacute patients with stroke when combined with trunk restraints. The modified therapy was found to be more effective than the traditional form and the inclusion of the trunk constraint enabled this therapy form to be more effective.

Several other studies have investigated different manual therapy approaches with more specific conditions. Kaya Mutlu et al. compared the effects of two manual techniques (i.e., passive joint mobilization and mobilization with movements) and a modality (i.e., electrotherapy) to determine which is more effective in treating patients with knee osteoarthritis^22^. The authors found that patients whose interventions included a manual therapy technique experienced a larger reduction in pain and demonstrated significant improvement in all outcome measures in the short and long-term compared to those who underwent electrotherapy^22^. In other studies that compared two to three different therapy techniques, the results found that either both or all techniques had a similar^23^ or negligible^24,25^effect. Burke et al. compared two manual therapy interventions (i.e., Graston instrumented-assisted soft tissue mobilization and soft tissue mobilization administered by a clinician) to evaluate their effectiveness in treating carpal tunnel syndrome^23^. The authors found that after both therapy interventions, significant improvements were shown in total wrist strength, wrist motion, and median nerve conduction latencies. This finding indicated that both manual therapy techniques had equivalent results.

In a later study by Cleland et al., the authors evaluated three manual physical therapy techniques (supine thrust manipulation, side-lying thrust manipulation, and non-thrust manipulation) in a group of patients with low back pain. The authors found significant differences in the Numerical Pain Rating Scale (NPRS) and Oswestry Disability Questionnaire (ODQ) when comparing the non-thrust group with the thrust groups during weeks 1 and 4 and 6 months^24^. At follow-up time points, both thrust groups had significantly lower ODQ scores compared to the non-thrust group. During weeks 1 and 4 both thrust groups had significantly lower NPRS scores compared to the non-thrust group. It is important to note that at the 6-month mark, all three interventions were equally effective in lowering NPRS scores. Furthermore, a study conducted by Pupin et al. compared the effects of two respiratory therapy techniques (i.e., Expiratory Flow Increase Technique [EFIT] and Postural Drainage [PD] - vibration accompanied) in infants with acute viral bronchiolitis^25^. The authors found no significant differences in the EFIT and PD interventions when evaluating their effect on the infant’s respiratory rate, heart rate, and oxygen saturation levels. However, the authors suggested that respiratory physical therapy was beneficial for the infants over time compared to no treatment.

Thus far, research has focused on comparing different manual therapy technique approaches with specific medical conditions. These comparison studies have been able to identify techniques that demonstrate the best clinical outcomes when treating patients. Although many changes in manual techniques have taken place, there are no available studies or published data that have systematically identified the changes in these universally used methods in physical therapy. It is important to identify and examine these changes to understand how manual therapy techniques have revolutionized the therapy field. This approach will help identify the most frequently used manual therapy techniques and how these have remained effective over time.

Thus, this study aimed to evaluate and analyze the changes in manual therapy techniques that have occurred over time in physical therapy through the following research questions: 1) how manual therapy techniques used by retired and currently licensed Washington state physical therapists have changed over time and 2) what are the most frequently used manual therapy techniques. These research questions examined how the use of manual therapy techniques has evolved from the perspective of physical therapists.

## Methodology

### Participants

Eighteen current and retired physical therapists, who currently practice or previously practiced in Washington state participated in this study. We excluded physical therapists licensed in other states because of the differences in state laws in relation to what manual therapy techniques can be practiced. Participants had between 1 – 44 years of experience working as licensed physical therapists in Washington state.

This study was approved by the Institutional Review Board (IRB) at Washington State University. Physical therapists were recruited from Whitman Hospital and Medical Clinics in Colfax, as well as from clinics and hospitals in Pullman, Yakima, Seattle, Spokane, Wenatchee, and other surrounding cities in Washington state. A recruitment email was sent to potential participants in clinics and hospitals upon receiving site approval. Participants were also recruited through the list serve of the APTA Washington, a Chapter of the American Physical Therapy Association, and previously established professional connections. In addition, recruitment posts were created and shared through various social media platforms such as Facebook, LinkedIn, and Twitter. Participants completed an online consent form and the Physical Therapy Profile Questionnaire^26^ hosted on Qualtrics, (Qualtrics, Provo, UT, USA) at their convenience.

### Assessment

A modified version of the Physical Therapy Profile Questionnaire^26^ was used. The PTPQ is a survey that is used to gather information from physical therapists, including (A) general information (e.g., education and demographics), (B) practice profile (e.g., years in practice, the current area of practice, types of roles performed in practice, and types of modalities performed before performing manual therapies), (C) treatment preferences (e.g., types of treatment approaches often used and/or recommended in practice), and D) practice changes (e.g., frequently used manual techniques, experienced manual techniques changes, and reasons for changing manual techniques). We modified the original version of the PTPQ to include an updated list of old vs. new manual techniques for physical therapists to identify the treatment approaches used in their practice and incorporated a set of new questions tailored to our research questions. Thus, the PTPQ enabled us to evaluate why there were changes in manual therapy techniques, how the changes affected patient outcomes, and identify preferred manual therapy techniques. Importantly, this questionnaire allowed the examination of how the field of physical therapy has evolved from the perspective of physical therapists.

### Data Analysis

A mixed methods approach was used to assess the reported preferred choices and changes in the use of manual techniques. To identify the most used and recommended manual therapy techniques, we used a frequency count to determine the total number of selections for each technique and then ranked them from highest to lowest based on the counts. We calculated the average frequency to identify the manual therapy techniques that have changed the most and their associated effect on patient outcomes using a 4-point rating scale. Additionally, participants were asked, “What was the reason for the change(s) in manual therapy technique(s)?” to identify why physical therapists changed their techniques. Participants were also asked, “What have been some notable changes in manual therapy techniques that have taken place during your years of practice?” to evaluate the experiences of physical therapists with manual therapy techniques over time. We analyzed these qualitative responses using a thematic network approach^27^ that organizes qualitative responses into global themes.

To identify whether there was a relationship between experience and recommended manual therapy techniques, we conducted a Pearson correlation between the most used manual therapy techniques and years of practice. We used SPSS Version 27.0 (IBM Corp., Armonk, NY, USA) to conduct all statistical analyses.

## Results

### Participant Demographics

Eighteen participants completed the PTPQ investigating the changes in manual therapy techniques in physical therapy. We excluded five participants because they did not meet the inclusion criteria and/or did not complete the consent form. Thus, the results reported here reflect data from the 13 participants who met the inclusion criteria. Of these 13 participants, 12 participants (nine males and three females) are currently licensed physical therapists in Washington state and one participant (female) is a retired and previously licensed physical therapist in Washington state. The mean age of the participants was 41±12.4 (range: 26 – 67) years. The mean years of practice were 14.5±13.4 (range: 1 – 44) years. Two participants had a bachelor’s degree, one participant had a master’s degree, nine participants had a Doctor of Physical Therapy degree, and one participant did not specify their education level. In addition to being licensed in Washington state, four participants were currently or previously licensed in another state, and two participants were licensed in two other states. We asked participants to provide their current areas of practice (e.g., general practice, geriatric/neurologic, education, sport/musculoskeletal, pediatric therapy, cardiopulmonary rehabilitation, wellness/health promotion, community development, research). The most common areas of practice were orthopedics with 12 selections, followed by sports with six selections, and geriatrics and general practice with three selections each. Three participants reported working in one area of practice, four participants reported working in two areas of practice, and six participants reported working in three or more areas of practice. The work environment also varied for participants, as three participants reported working in an outpatient hospital-based clinic, eight participants reported working in an outpatient private clinic, one participant reported working in acute care, and one participant reported working in a university or college. See Table 1 for detailed demographic information about the participants.

**Table 1:**
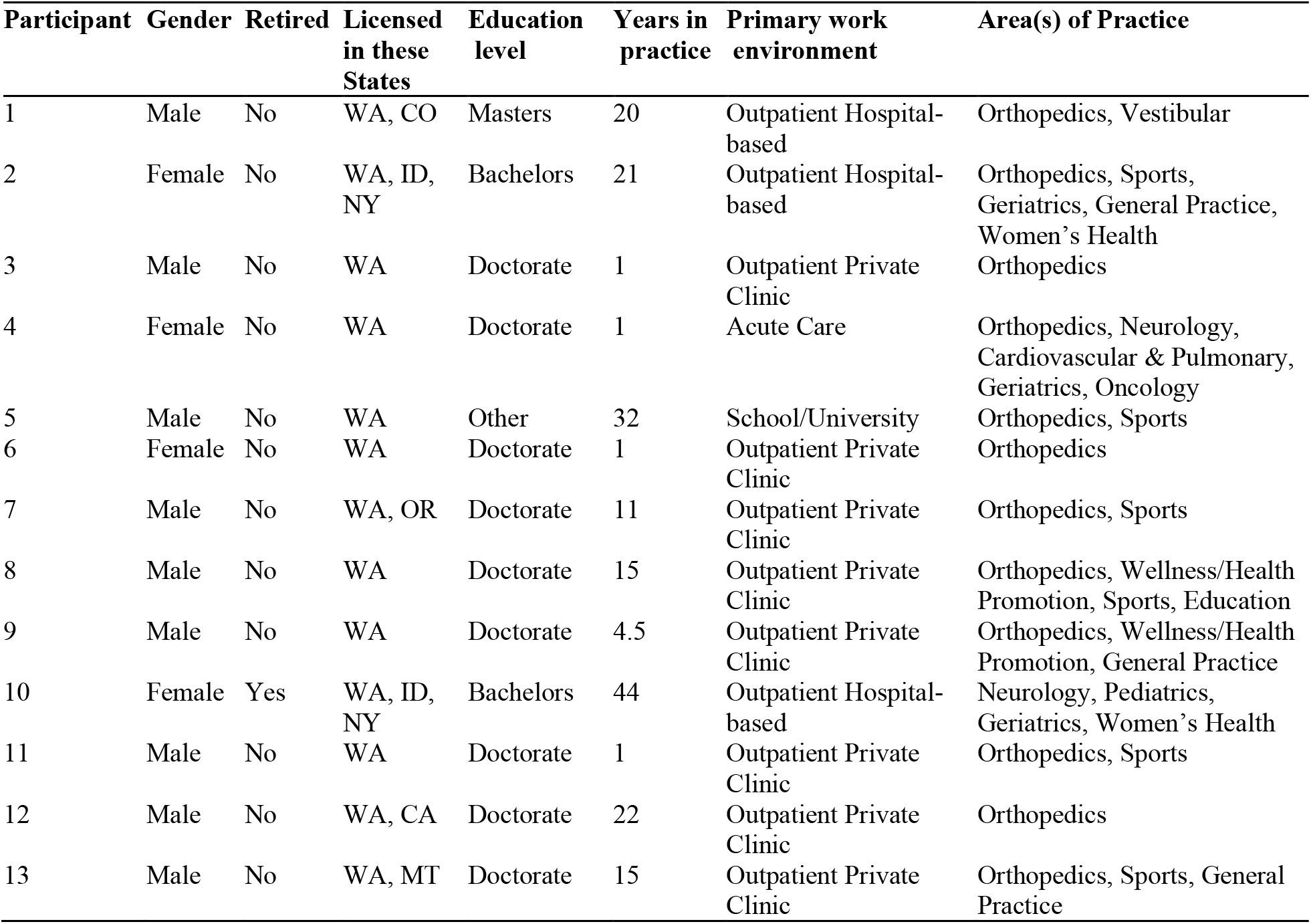
Characteristics of participants.

| Participant | Gender | Retired | Licensed in these States | Education level | Years in practice | Primary work environment | Area(s) of Practice |
| --- | --- | --- | --- | --- | --- | --- | --- |
| 1 | Male | No | WA, CO | Masters | 20 | Outpatient Hospital-based | Orthopedics, Vestibular |
| 2 | Female | No | WA, ID, NY | Bachelors | 21 | Outpatient Hospital-based | Orthopedics, Sports, Geriatrics, General Practice, Women's Health |
| 3 | Male | No | WA | Doctorate | 1 | Outpatient Private Clinic | Orthopedics |
| 4 | Female | No | WA | Doctorate | 1 | Acute Care | Orthopedics, Neurology, Cardiovascular & Pulmonary, Geriatrics, Oncology |
| 5 | Male | No | WA | Other | 32 | School/University | Orthopedics, Sports |
| 6 | Female | No | WA | Doctorate | 1 | Outpatient Private Clinic | Orthopedics |
| 7 | Male | No | WA, OR | Doctorate | 11 | Outpatient Private Clinic | Orthopedics, Sports |
| 8 | Male | No | WA | Doctorate | 15 | Outpatient Private Clinic | Orthopedics, Wellness/Health Promotion, Sports, Education |
| 9 | Male | No | WA | Doctorate | 4.5 | Outpatient Private Clinic | Orthopedics, Wellness/Health Promotion, General Practice |
| 10 | Female | Yes | WA, ID, NY | Bachelors | 44 | Outpatient Hospital-based | Neurology, Pediatrics, Geriatrics, Women's Health |
| 11 | Male | No | WA | Doctorate | 1 | Outpatient Private Clinic | Orthopedics, Sports |
| 12 | Male | No | WA, CA | Doctorate | 22 | Outpatient Private Clinic | Orthopedics |
| 13 | Male | No | WA, MT | Doctorate | 15 | Outpatient Private Clinic | Orthopedics, Sports, General Practice |

### Frequently used manual therapy techniques

We asked participants, “Do/Did you perform a modality (E-STIM, Ultrasound, Heat/Ice) on a patient before performing manual therapy techniques?” Ten participants reported that they have not used a modality before performing manual therapy on a patient, whereas three participants did report the usage of a modality (i.e., heat, ultrasound, bike, and/or treadmill) before performing manual therapy on a patient.

In addition, we asked participants to identify their most used and recommended manual therapy technique(s) in their practice from the following list: soft tissue mobilization techniques, muscle energy techniques, positional changes, Mulligan’s technique, myofascial release, positional techniques, joint mobilization, joint manipulation, dry needling, cupping, augmented soft tissue manipulation, instrument assisted soft tissue mobilization, and taping. Joint mobilization was the most used manual therapy technique with 11 indications of usage, followed by taping with 10 indications, and muscle energy technique with nine indications (see Figure 1). Of all the manual therapy techniques listed, dry needling was the only technique not selected by any of the participants. This is not surprising as state law prohibits physical therapists from performing dry needling in six states including Washington, but it is legal in 36 states including D.C.

**Figure 1:**
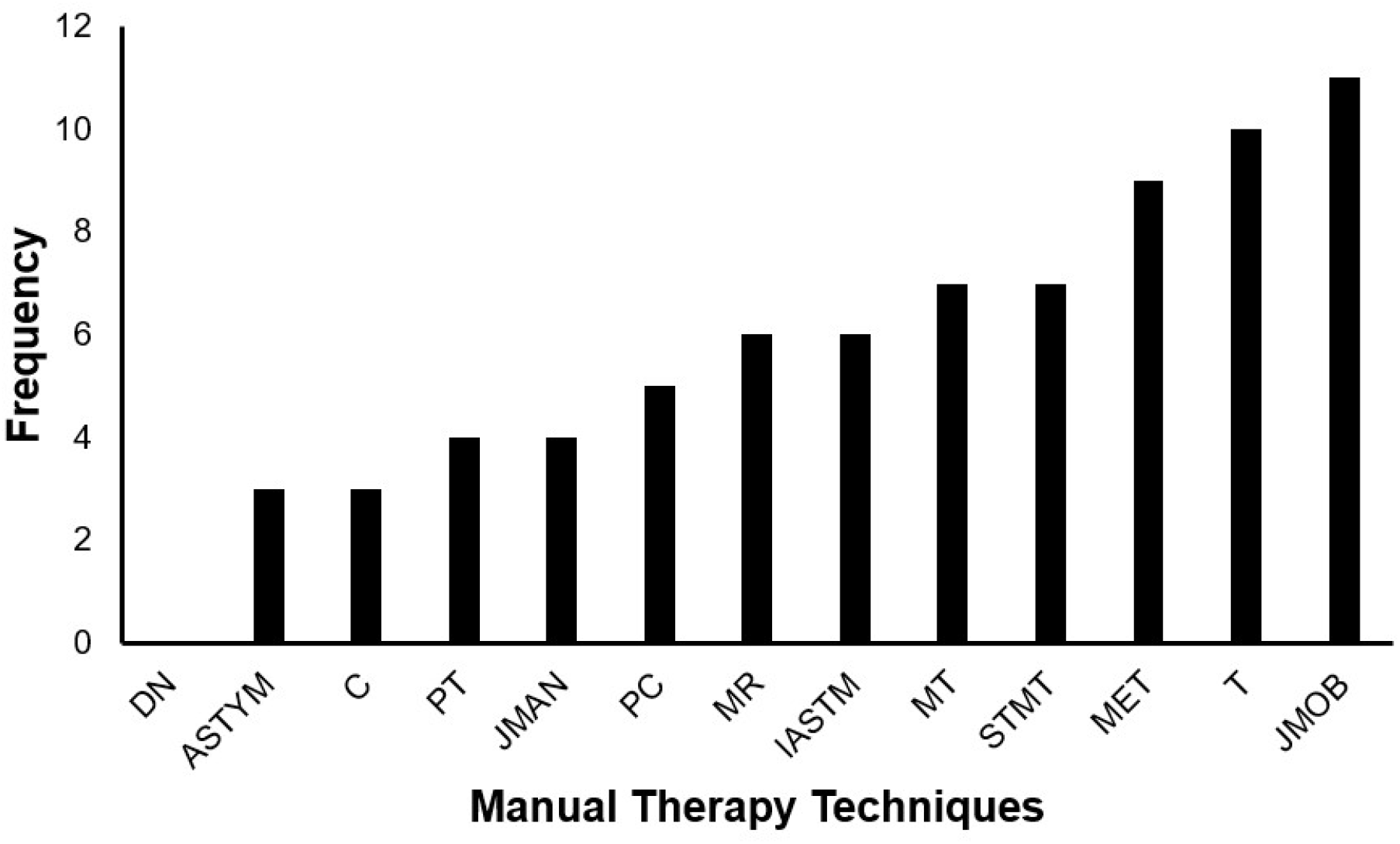
Frequency of Manual Therapy Techniques from the least to highest used or recommended. DN, Dry Needling; ASTYM, Augmented Soft Tissue Manipulation; C, Cupping; PT, Positional Techniques; JMAN, Joint Manipulation; PC, Positional Changes; MR, Myofascial Release; IASTM, Instrument Assisted Soft Tissue Mobilization; MT, Mulligan’s Technique; STMT, Soft Tissue Mobilization Techniques; MET, Muscle Energy Techniques; T, Taping; JMOB, Joint Mobilization.

### Changes in the use of manual therapy techniques

We asked participants about changes in their practice, specifically if they have changed their use of any of the manual therapy technique(s) over time. Eight participants indicated that they have changed their use, while five participants indicated that they did not. For those eight participants that indicated changes, we asked the following questions, “If yes, what manual therapy technique(s) have/did you change(d)? Was there a positive effect on the patient outcome?” The participants were asked to select the manual therapy technique(s) they changed and rated them on a 4-point scale for their associated effect on patient outcome, where 1 = No Effect; 2 = Little Effect; 3 = Somewhat of an Effect; and 4 = Big Effect (i.e., whether the change in the use of the technique has no effect, little effect, somewhat of an effect, or big effect on patient outcome; see Figure 2). Myofascial release was reported as the most changed manual therapy technique with a total rating of 16. Soft tissue mobilization technique was the second most changed technique with a total rating of 13. Mulligan’s technique and instrument-assisted soft tissue mobilization were both identified as the third most changed techniques with total scores of 8 each. We further asked these eight participants to report the reason they made changes in the manual therapy techniques. The most common reasons for changing their manual therapy techniques were due to the amount of experience of the physical therapist in the field. One participant, who had 11 years of practice stated, “Over time, I learned when to do what technique at the right time for the right person.”

**Figure 2:**
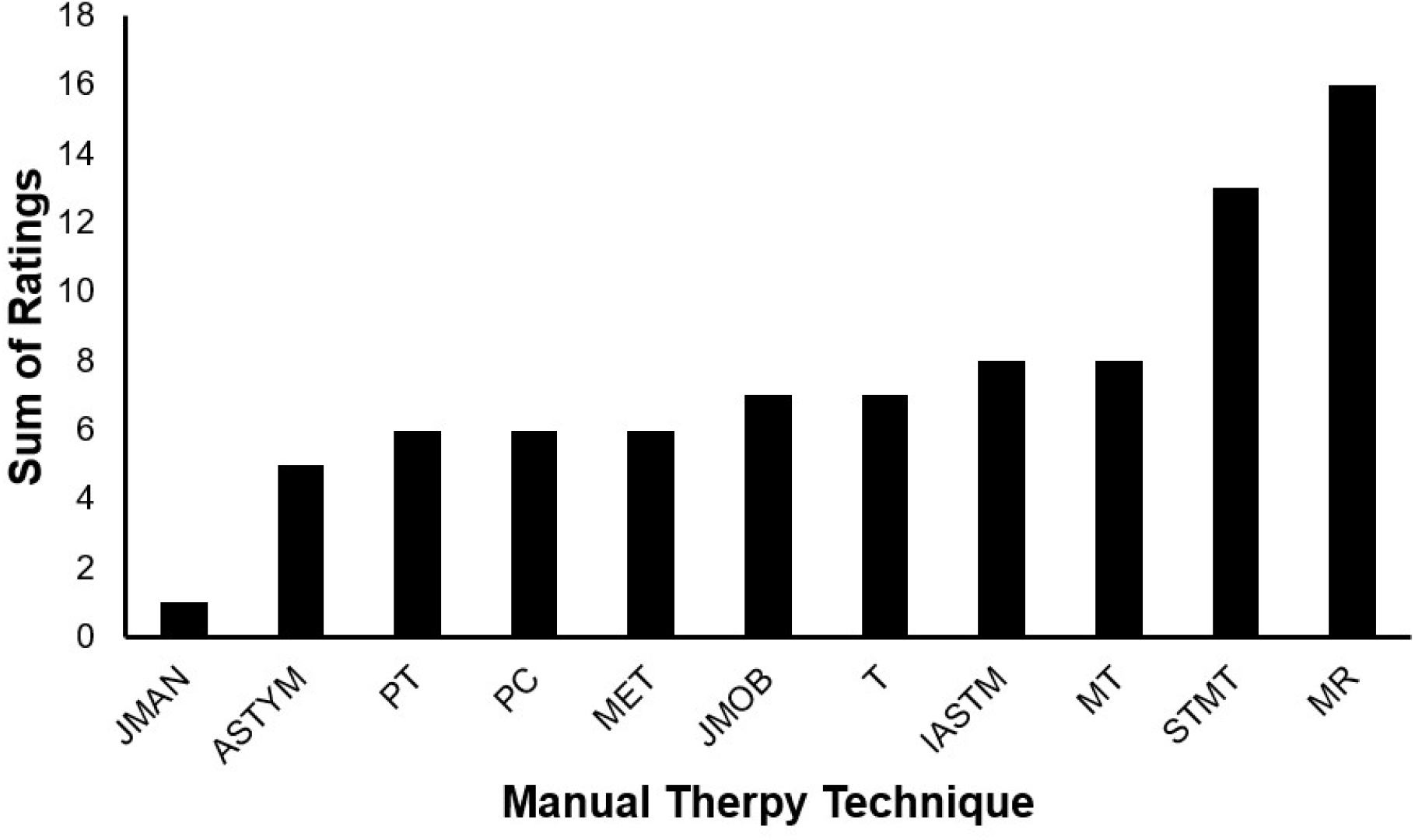
Manual Therapy Techniques and Their Sum of Ratings Based on Their Changes. JMAN, Joint Manipulation; ASTYM, Augmented Soft Tissue Manipulation; PT, Positional Techniques; PC, Positional Changes; MET, Muscle Energy Techniques; JMOB, Joint Mobilization; T, Taping; IASTM, Instrument Assisted Soft Tissue Mobilization; MT, Mulligan’s Technique; STMT, Soft Tissue Mobilization techniques; MR, Myofascial Release.

Lastly, we asked participants, “What have been some notable changes in manual therapy technique(s) that have taken place during your years in practice?” The responses to this question varied among the eight participants, but there were two common themes. One theme was that there has been a significant increase in manual techniques and tools that are helping improve the physical therapy field and the second theme was having experienced a significant decrease in usage and teaching of manual therapy techniques. One participant, with 21 years of experience, stated, “Fewer younger therapists perform them or were even taught manual techniques in school. A big disservice to our patients and profession in my opinion.” Participants also commented on the increased availability of techniques and another participant, with 44 years of experience, stated, “Manual therapy became available in my senior year of PT school. Far more techniques now.”

### Relationship between experience and use of manual therapy techniques

We correlated the total years of practice with participants’ preferred choices of manual techniques (see Figure 3). There was no significant correlation between the years of practice and the number of manual therapy techniques used by participants (*r* = -0.044, *p* = 0.887).

**Figure 3:**
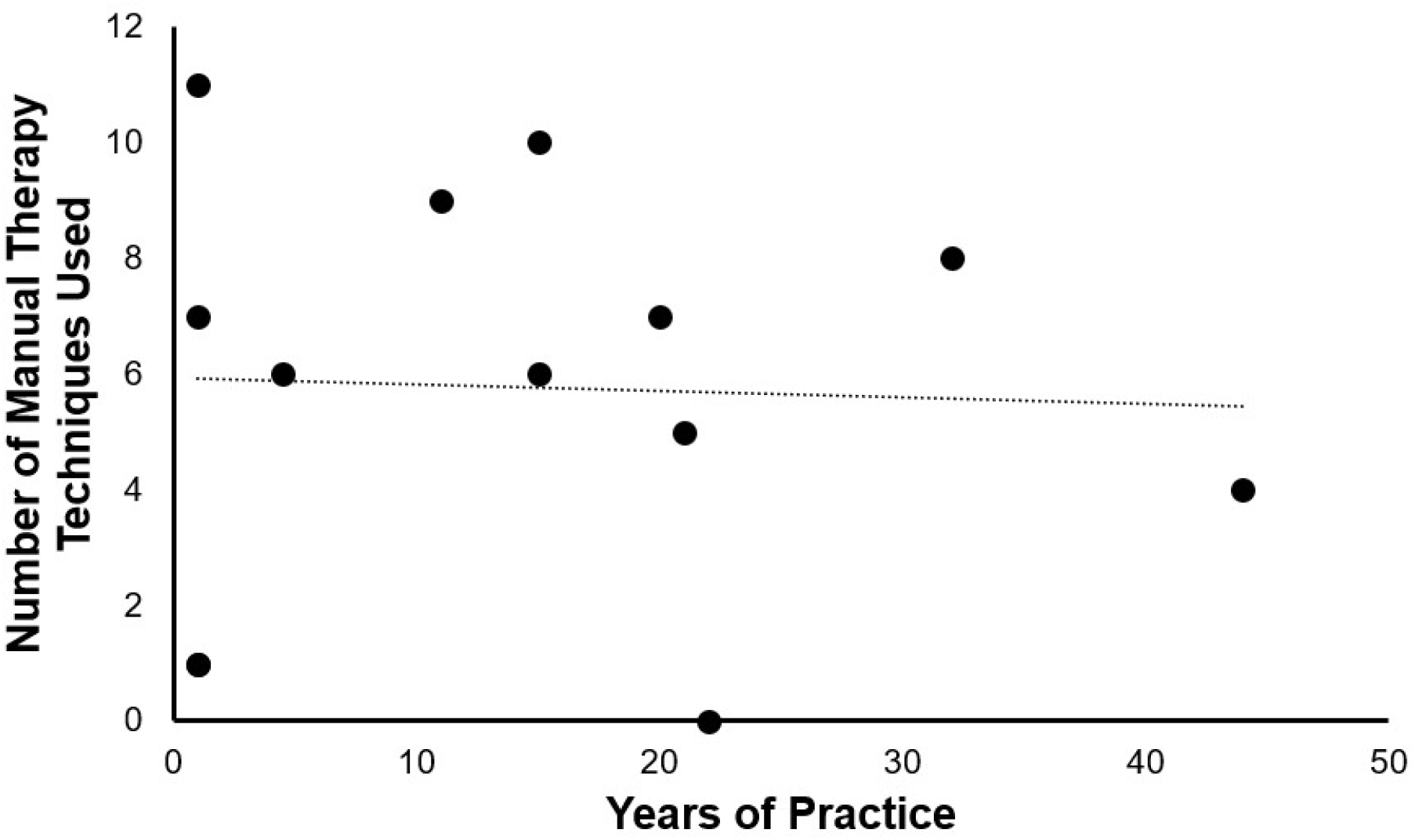
Correlation between years of practice and number of manual therapy techniques used or recommended.

We further evaluated the four physical therapists with one year of experience, two of whom reported high usage of manual therapy techniques (participants 3 and 6) and two who reported a low usage of manual therapy techniques (participants 4 and 11). Participant 3 reported practicing in one area (i.e., orthopedics), and reported using seven manual therapy techniques (i.e., soft tissue mobilization techniques, muscle energy technique, positional changes, Mulligan’s technique, joint mobilization, instrument-assisted soft tissue mobilization, and taping). Participant 6 reported practicing in two areas (i.e., orthopedics and sports), and reported using 11 manual therapy techniques (i.e., soft tissue mobilization techniques, muscle energy techniques, positional changes, Mulligan’s technique, myofascial release, positional techniques, joint mobilization, joint manipulation, cupping, instrument assisted soft tissue mobilization, and taping). Participant 4 reported practicing in five areas (i.e., orthopedics, neurology, cardiovascular and pulmonary, geriatrics, and oncology), but only reported using one manual therapy technique (i.e., taping). Participant 11 reported practicing in two areas (i.e., orthopedics and sports), but only reported using one manual therapy technique (i.e., joint mobilization).

We then evaluated the remaining nine participants, who had four or more years of practice. These participants reported using an average of 6.1 manual therapy techniques and engaging in two or more areas of practice. The exception to this group was participant 12 with 22 years of experience, who reported one area of practice and did not report any usage of manual therapy techniques.

## Discussion

The primary purpose of this study was to investigate changes in manual therapy techniques in physical therapy from the perspective of physical therapists in Washington state. Specifically, we recorded the most frequently used manual therapy techniques and asked participants how the use of these techniques had changed over time from their perspective. We found that the most commonly used techniques were joint mobilization, taping, and muscle energy techniques. The usage of these manual therapy techniques changed based on the experiences of physical therapists to improve patient outcomes.

### Top three most frequently used techniques

The top three most frequently used manual therapy techniques were joint mobilization, taping, and muscle energy technique. These results are surprising, as other studies consider massage therapy (commonly referred to as soft tissue mobilization techniques) to be the most used and practiced manual therapy technique^4,5^. However, when assessing the historical evolution of these techniques, the most common manual therapy techniques found in this study have remained universally used since their initial origins and introduction to physical therapy. For example, in 400 BCE, Hippocrates described the setting of bones by a method of leverage which is a similar method to joint mobilization^5^. Furthermore, muscle energy technique is a manual technique that dates to 1948 and has been widely used in both osteopathic medicine and physical therapy as it improves musculoskeletal function^28^. Lastly, taping (commonly known as Kinesio Tape and/or McConnell taping) was invented in the 1970s by the Japanese chiropractor Dr. Kenso Kase, to help with stability, pain relief, injury prevention, muscle inhibition, and other common usages^29^. The long history of these three manual therapy techniques provides support for their common use by our participants.

We found no correlation between the years of practice and the number of manual therapy techniques used and recommended by physical therapists. This may be due to the amount of variability among participants. For example, physical therapists with one year of experience reported using a range of techniques from just one to 11. Compared to physical therapists with one year of experience, physical therapists with more than 4.5 years of experience reported using a range of techniques from 5-10. As the sample size in the present study is small, this variability may be concealing relationships between experience and the number of techniques used. These results may also be influenced by the geographic locations of where participants conducted their practice. This may influence the types of populations and the variability of conditions they treat. In addition, the reported amounts of techniques used could be influenced by the primary work area of the physical therapists. Participants were not asked to specify which reported areas of practice were current versus past. Therefore, we could not determine if changes in the number of manual techniques reported were related to a change in a specific area of practice. In future studies, it would be beneficial to have data about their specific area of work to then compare with the number of reported manual therapy techniques. Another aspect to consider is the available types of therapy equipment and/or modalities at each facility that could influence the number of therapy techniques being used by the physical therapist.

### Changes in use of manual therapy techniques

Our results suggest that the use of manual therapy techniques has changed over time because of the individual experience of each physical therapist as they continue to improve outcomes for patients. The modification in techniques stems from the experience of working with individuals of diverse backgrounds and learning which manual therapy techniques work best for that individual and/or the specific injury. These results align with the findings of earlier studies, such as the one by Kaya Mutlu et al. that compared two manual therapy techniques (i.e., mobilization with movements and passive joint mobilization) along with a modality (i.e., electrotherapy) to evaluate the effectiveness in treating two groups of patients with knee osteoarthritis^22^. The authors found that participants who underwent the manual therapy had a significant decrease in pain and had better ratings in their outcome measures compared to the group who underwent electrotherapy. These findings suggest that physical therapists make adjustments by identifying which technique has the best outcome for a patient.

Furthermore, our results suggested differences in perspective on changes in manual therapy techniques. Some physical therapists reported that there has been a significant increase in the usage of manual therapy techniques and tools that have helped improve the physical therapy field, whereas other physical therapists experienced a decrease in the usage of manual therapy techniques. These contrasting views may reflect a variety of factors, including differences in training at different physical therapy programs, availability of modality instruments, and experience with different types of populations that should be investigated in future studies.

### Factors that may influence the use of manual therapy techniques

The PTPQ did not ask the participants to specify the geographic location and demographics of patients in their area of work. These factors may influence the manual therapy techniques participants used because different geographic locations will contain their diverse population of individuals from different professional backgrounds, thus injuries, medical conditions, and accidents will vary based on the type of jobs or vocations within those geographic locations. Some geographic locations may be high in industrial jobs, compared to rural areas where farming and agriculture are the main source of jobs, creating variability in the types of injuries physical therapists manage. In addition to the variability of jobs in the different geographic areas, the variability in population demographics within each geographic area will also impact the types of manual therapy techniques used based on the unique injuries from performing a certain sport, activity, or task. This is a key area to investigate as there is currently no research available that has investigated how different geographic and demographic areas influence and impact the usage of manual therapy techniques.

Geographic location not only impacts the patient demographics, injuries, or disorders that physical therapists manage, but different states also have different laws related to what manual therapy techniques physical therapists can practice. For example, the law prohibits physical therapists from performing dry needling in Washington state as well as in five other states, but it is legal in 36 states in the United States. Thus, future studies should focus on expanding recruitment to include physical therapists from other states to assess how the usage of manual therapy techniques and modalities changes with state regulations.

In addition, another factor to consider is the availability of different modalities at each clinic and how this impacts the usage of manual therapy techniques. Given the costly prices of new modalities, not all clinics are able to afford them and provide them to their clients. For example, physical therapists who practice in a small private clinic may not have access to the same equipment as those practicing in hospital-based clinics. This limits physical therapists to utilize a variety of manual therapy techniques compared to those practices that can afford more expensive equipment and may rely on them rather than manual techniques. Thus, some physical therapists may utilize modalities to treat patients and even eliminate some of the manual therapy work if this form of therapy is more effective.

Furthermore, the curriculum followed by different physical therapy programs and manual therapies learned by the physical therapist from their institutions can lead to differences in practice. For example, some physical therapy programs emphasize the exposure of students to clinical internships or other opportunities within the second semester of their program. This allows students to have hands-on experience at an early stage of the program compared to other programs that do not introduce these experiences until the last year. The early exposure to clinical practice allows students to not only gain experience but allows for opportunities to gain experience from instructors and clinicians to learn more manual therapy techniques. This information can help provide context for the different learned manual therapy techniques that each participant was exposed to through their educational curriculum.

### Limitations and future studies

The results of this study are limited by the small sample size and limited geographical location of participants. A larger sample size will allow for the expansion of the current findings. We faced obstacles when collecting the data due to unclear responses that lacked depth and/or missing responses to questions. These limitations can be overcome by interviewing participants in person or through teleconferencing software to collect more detailed responses. In addition, future studies can collect data on the institutions from which participants obtained their degrees and whether there are differences in the curriculum that can impact the practice of physical therapy.

This study was also limited in the quantity and types of questions we were able to ask the participants, as well as the different areas we wanted to investigate due to time restraints. We took into consideration the amount of time we were asking of participants since the study had no compensation for participation and the limited time participants have during their work schedule since most physical therapists meet with over 15 clients per day. Nonetheless, the results of this study serve as a stepping-stone to further investigate how manual therapy techniques are evolving and changing the field of physical therapy from the perspective of physical therapists.

## Conclusion

While previous research has focused on comparing the effects and efficacy of different manual therapy techniques, the results of this study examined changes in manual therapy techniques from the perspective of physical therapists. Our results suggest that the changes in manual therapy techniques have been due to the progression of experience of physical therapists. More specifically, learning when to perform a certain manual therapy technique with a patient at the appropriate time, can result in a better patient outcome. In addition, from the personal views of the eight physical therapists that reported changes throughout their careers, about half of them reported experiencing positive improvements. This arose from learning new manual techniques and/or acquiring new assistive tools to help treat patients and improve patient outcomes. This study poses new insights on the usage and change in manual therapy techniques in physical therapy and serves as a stepping-stone to further investigate how manual therapy techniques are evolving and changing the field of physical therapy from the perspective of physical therapists.

## Data Availability

All data produced in the present study are available upon reasonable request to the authors.

